# Efficacy and Safety of Leflunomide for Refractory COVID-19: An Open-label Controlled Study

**DOI:** 10.1101/2020.05.29.20114223

**Authors:** Qiang Wang, Haipeng Guo, Yu Li, Xiangdong Jian, Xinguo Hou, Ning Zhong, Jianchun Fei, Dezhen Su, Zhouyan Bian, Yi Zhang, Yingying Hu, Yan Sun, Xueyuan Yu, Yuan Li, Bei Jiang, Yan Li, Fengping Qin, Yingying Wu, Yanxia Gao, Zhao Hu

**Affiliations:** Department of Nephrology, Qilu Hospital (Qingdao), Cheeloo College of Medicine, Shandong University, Qingdao, China; Department of Critical Care Medicine, Qilu Hospital, Cheeloo College of Medicine, Shandong University, Jinan, China; Department of Respirology, Qilu Hospital, Cheeloo College of Medicine, Shandong University, Jinan, China; Department of Emergency Medicine, Qilu Hospital, Cheeloo College of Medicine, Shandong University, Jinan, China; Department of Endocrinology, Qilu Hospital, Cheeloo College of Medicine, Shandong University, Jinan, China; Department of Gastroenterology, Qilu Hospital, Cheeloo College of Medicine, Shandong University, Jinan, China; Department of Anesthesiology, Qilu Hospital, Cheeloo College of Medicine, Shandong University, Jinan, China; Department of Psychiatry, Renmin Hospital of Wuhan University, Wuhan, China; Department of Quality control, Qilu Hospital, Cheeloo College of Medicine, Shandong University, Jinan, China; Department of Nephropathy, Shandong Provincial Third Hospital, Cheeloo College of Medicine, Shandong University, Jinan, China; Department of Nephropathy, Qilu Hospital, Cheeloo College of Medicine, Shandong University, Jinan, China

## Abstract

**OBJECTIVE:** To evaluate the safety and efficacy of leflunomide for the treatment of refractory COVID-19 in adult patients.

**DESIGN:** Open-label controlled study

**SETTING:** A designated hospital for patients with refractory COVID-19 in Wuhan, China.

**PARTICIPANTS:** 27 hospitalized adult patients (≥18 years of age) with radiologically confirmed pneumonia and SARS-CoV-2 positive for more than 28 days despite standard care were assigned to receive standard of care (SOC, grp I) or leflunomide + SOC (grp 2). After 2 weeks, grp 1 and grp 2 patients who continued to be SARS-CoV-2-positive received leflunomide for 14 days while continuing SOC.

**MAIN OUTCOME MEASURES:** The primary outcomes were the rate of and time to SARS-CoV-2 clearance and the 14-day and 30-day hospital discharge rate.

**RESULTS:** Twelve patients enrolled in grp 1 and 15 patients were in grp 2. The 14 days SARS-CoV-2 viral clearance rate was 80.0% (12/15) for grp 2 patients receiving leflunomide *versus* 16.7% for grp 1 patients (2/12) (*P*=0.002). By day 14, the median time to SARS-CoV-2 clearance was 6.0 days (range 1-12, IQR 1-12) for grp 2 patients. In grp 1, two patients converted to viral negative on days 1 and 6 (*P*=0.002). The 14-day discharge rate was 73.3% (11/15) for the grp 2 *versus* 8.3% (1/12) for grp 1 (*P*=0.001). The 30-day discharge rate was 100% (15/15) for the grp 2 *versus* 66.7% (8/12) for grp 1. No severe adverse events or deaths were reported.

**CONCLUSION:** Leflunomide is effective in enhancing SARS-CoV-2 clearance and hospital discharge in refractory COVID-19 patients. The addition of leflunomide to SOC did not increase adverse events versus SOC. These preliminary observations underscore a need for a randomized clinical study of leflunomide in SARS-CoV-2 infection.

**WHAT IS ALREADY KNOWN ON THIS TOPIC:** Based on the large numbers of global infected patients of SARS-CoV-2, there will be many patients on persist viral positive which is named refractory covid-19. Specific medication for the treatment of the refractory covid-19 has been approved.

Leflunomide has been widely used in rheumatoid arthritis and psoriatic arthritis with good safety and tolerance. Recently, it is found an activity of anti-SARS-CoV-2 in vitro and the effective concentration of leflunomide is within the recognized therapeutic level for rheumatoid arthritis.

**WHAT THIS STUDY ADDS:** Leflunomide is effective in enhancing SARS-CoV-2 clearance and hospital discharge in refractory COVID-19 patients. The safety and tolerability of the 14-28-day course of treatment with leflunomide is acceptable.

## Introduction

Coronavirus disease 2019 (COVID-19) is a respiratory infectious disease caused by the severe acute respiratory syndrome coronavirus 2 (SARS-CoV-2). SARS-CoV-2 pandemic has so far spread to 210 countries and infected approximately three million people globally and caused more than 200,000 deaths^1^. Even though the majority of patients with COVID-19 clear SARS-CoV-2 within 14-20 days, our observations indicate that SARS-CoV-2 persists in 2-5% of infected hospitalized patients up to 4 weeks despite current best available care^2-6^. These patients are defined as having refractory or chronic COVID-19, and are not discharged from the hospital due to the risk of shedding the virus in communities.

Given the current lack of effective treatment for COVID-19, repurposing existing drugs to treat COVID-19 offers a rational option to tackle this global public health emergency^7^. Leflunomide is an isoxazole derivate with immunosuppressive activities and has been used as disease-modifying anti-rheumatic drugs in rheumatoid arthritis and psoriatic arthritis^8-12^. The drug also possesses anti-viral activities against BK polyomavirus and cytomegalovirus^13-18^. Schläpfer et al. reported that leflunomide decreased HIV replication by approximately 75% at concentrations that can be obtained with conventional dosing^19^. Martin et al. observed a strong dose-dependent decrease in replication and transcription of the Ebola virus in the presence of teriflunomide (the active metabolite of leflunomide)^20^. Upon oral administration, over 80% of leflunomide is converted to teriflunomide via hepatic metabolism in the first pass through the liver^21,22^. At standard doses for rheumatoid arthritis, teriflunomide inhibits *de novo* pyrimidine synthesis *via* dihydroorotate dehydrogenase (DHODH), and at higher doses suppresses tyrosine and serine kinase activities^23^. Leflunomide is safe and well tolerated^9-10^. A recent study suggested that leflunomide/teriflunomide could be repurposed for SARS-CoV-2 therapy^24^ with a therapeutic range between 6-26 μM of teriflunomide for SARS-CoV-2, which is within the recognized therapeutic level for rheumatoid arthritis^8^.

In this non-randomized, controlled, open-label trial, we evaluated the safety and efficacy of leflunomide for refractory COVID-19 in adult patients who were hospitalized between March 13 and April 17 of 2020 at Hu Bei Renmin Hospital in Wu Han City, China. Crossover of control patients to leflunomide treatment was allowed if control patients were still SARS CoV-2 positive after 14 days in the control arm.

## Methods

### Study Design

This open-label study of leflunomide as adjunctive therapy to SOC was designed as a pilot study in anticipation for a randomized controlled trial (ChiCTR2000030058) and enrolled hospitalized adult patients (≥ 18 years of age) with radiologically confirmed COVID-19 pneumonia who were reverse transcriptase-polymerase chain reaction (RT-PCR)-positive for SARS-CoV-2 for more than 28 days despite standard care. Eligible patients had pneumonia confirmed by chest imaging and had an oxygen saturation (SaO2) of 94% or higher on room air PaO2/FiO2 ratio ≥ 300 mg Hg (ratio of the partial pressure of oxygen (PaO2) to the fraction of inspired oxygen (FiO2)). Exclusion criteria were pregnancy, a history of liver disease, alanine aminotransferase level 5 times higher than the upper normal limit (50 U/L), and stage 4 chronic kidney disease.

The study protocol adhered to the SPIRIT statement^25^ and conducted under the International Conference on Harmonization Guidelines for Good Clinical Practice and the Declaration of Helsinki and the reporting of the study adhered to the CONSORT statement^26^. The corresponding author was responsible for the study design. All the authors contributed to the analysis of the data. All patients provided written informed consent to the study.

### Study Intervention

Patients were assigned to the standard care group (grp 1) or the leflunomide group (grp 2) via patient choice. Standard care was provided to all patients according to the guideline^27^ including supplemental oxygen and supportive care, as well as concurrent therapy with hydroxychloroquine, interferon-α, anti-human immunodeficiency virus drugs (lopinavir/ritonavir) or anti-influenza drugs (arbidol, oseltamivir) was allowed. Leflunomide (Airuohua, manufactured by Changzheng-Cinkate) was given at 30 mg/day to patients who were less than 64 years old, and 20 mg/day to patients who were ≥ 65 years old. The treatment lasted for 14 days as the first phase. All patients who continued to be RT-PCR-positive for SARS-CoV-2 by day 14 received leflunomide for 14 days as the second phase. Once RT-PCR for SARS-CoV-2 tests were negative for two consecutive assays over 24 hours apart, the physician could terminate the treatment and observe for two days before discharge.

### SARS-CoV-2 Testing

Throat swap samples were obtained by skilled nurses, and sputum samples were collected after patients were trained by nurses one day before and daily after the start of leflunomide therapy until discharge from the hospital. RNA was extracted using the MagNA Pure 96 system and semiquantitative real-time RT-PCR was performed on ABI 7500 PCR analyzer (Thermofisher) by using LightMix Modular SARS-CoV-2 (COVID-19) assays (TIB MOBIOL) with primers targeting the nucleocapsid protein (NP) gene and open reading frame (ORF) of SARS-CoV-2. A cycle threshold (C_t_) ≤ 38.0 was considered positive. The report was presented as a grade scale of 1+ ~ 3+ to indicate increasing viral load^2^. Two consecutive negative RT-PCR results 24 hours apart for SARS-CoV-2 indicated SARS-CoV-2 clearance.

### Study Outcomes

The primary outcomes included the rate of and time to SARS-CoV-2 clearance, and the 14-day and 30-day hospital discharge rate. Secondary outcomes included the incidence of flares, defined as the event of conversion to being RT-PCR-positive for SARS-CoV-2 after turning negative for SARS-CoV-2 in the course of treatment, and adverse events. Patients were discharged when clinical symptoms and radiographic images of COVID-19 were significantly improved and RT-PCR-negative for SARS-CoV-2 with two consecutive assays over 24 hours apart.

Adverse events (AEs) were graded and recorded according to NCI-CTCAE version 4.03. Safety events included AEs and severe adverse events (SAEs). SAEs included any untoward medical occurrence that resulted in death, was life-threatening, required hospitalization or prolongation of hospitalization, or caused significant or persistent disability or incapacity, or birth defects. AEs were coded to a preferred term using the Medical Dictionary for Regulatory Activities (MedDRA) 22.0. Safety assessments were based mainly on the occurrence, frequency, and severity of AE and analyzed mainly using descriptive statistics.

### Statistical Analysis

The analysis population included all patients. Analyses were descriptive in nature. Summary tabulations included the number of observations; mean, standard deviation, median, interquartile range (IQR), minimum and maximum for continuous variables; number and percentage per category for categorical data. The rate of SARS-CoV-2 clearance and discharge were analyzed by Fisher’s Exact Test. Time to SARS-CoV-2 clearance and time to discharge from the hospital were described with Kaplan-Meier analysis. All analyses were conducted with SPSS software, version 25 (IBM Corp.). *P*<0.05 indicated a statistically significant difference.

## Results

### Patient Demographic and Baseline Characteristics

The study flowchart is shown in Figure 1. 27 patients were enrolled in the study. Their median age was 62 (IQR 43-70), and 52% of the patients were men. Patient demographic and baseline characteristics are shown in Table 1. The median duration of symptomatic onset or positive SARS-CoV-2 was 45 days (IQR 41-50). Twelve (44%) patients on admission had severe COVID-19 pneumonia^28^. Sixty-seven percent of the patients had comorbidities. Upon enrollment, 15 patients were assigned to receive leflunomide (grp 2) and 12 patients to receive standard care only (grp 1). In grp 2, 10 patients received 20 mg/day leflunomide and 5 patients received 30 mg/day leflunomide. Except for gender, the two groups were comparable in the demographic and baseline characteristics. The median number of medicines received was 6 (IQR 5-8) in the leflunomide group and 7.5 (IQR 7-9) in the standard care group. Patient pharmacotherapy data are shown in Table S1.

**Figure 1.**
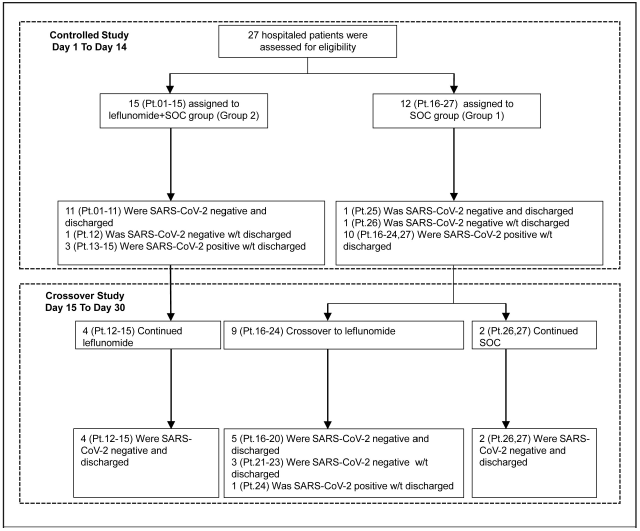
The Study Flowchart.

**Table 1.** Patient Demographic and Baseline Characteristics*.

| **Characteristic** | **Group 2**<br>**(Leflunomide+SOC)**<br>**(N = 15)** | **Group 1**<br>**(Standard of Care)**<br>**(N = 12)** | **Total**<br>**(N = 27)** |
| --- | --- | --- | --- |
| **Median age (IQR), years** | 65 (43-70) | 62 (46-69.5) | 62 (43-70) |
| <50 | 4 (27) | 3 (25) | 7 (26) |
| 50 to <70 | 6 (40) | 6 (50) | 12 (45) |
| ≥70 | 5 (33) | 3 (25) | 8 (29) |
| **Male sex** | 10 (67) | 4 (33) | 14 (52) |
| **Mean duration of symptom onset or**<br>**SARS-CoV-2 positivity before enrollment**<br>**(IQR), days** | 47 (44-49) | 42.5 (38-51.5) | 45 (41-50) |
| **Severe COVID-19 on admission** | 6 (40) | 6 (50) | 12 (44) |
| **Mild/Common COVID-19 on admission** | 9 (60) | 6 (50) | 15 (56) |
| **Clinical manifestations on admission** |  |  |  |
| Fever | 11 (73) | 10 (83) | 21 (78) |
| Dyspnea | 9 (60) | 7 (58) | 16 (59) |
| **Laboratory findings (IQR)** |  |  |  |
| Median ALT, IU/L | 23.5 (18-41.8) | 43 (24-77) | 26 (20.5-61.5) |
| Median AST, IU/L | 21 (18-35.3) | 34 (21-42) | 24 (18.5-35.5) |
| Median creatinine, μmol/L | 55 (50-71.8) | 54 (48-68) | 54 (50-69) |
| Median leucocyte count — ×10<sup>9</sup>/liter | 4.4 (3.7-6.3) | 5.1 (4.6-5.8) | 5.0 (4.2-6.1) |
| Median lymphocyte count — ×10<sup>9</sup>/liter | 1.6 (0.9-2.3) | 1.4 (1.1-1.7) | 1.4 (1.0-1.8) |
| **Comorbidities** |  |  |  |
| Any condition | 10 (67) | 8 (67) | 18 (67) |
| Hypertension | 4 (27) | 3 (25) | 7 (26) |
| Diabetes | 1 (7) | 1 (8) | 2 (7) |
| Hyperlipidemia | 4 (27) | 1 (8) | 5 (19) |
| Cardiovascular disease | 2 (13) | 1 (8) | 3 (11) |
| Cancer | 1 (7) | 0 | 1 (4) |

\*ALT denotes alanine aminotransferase, and AST aspartate aminotransferase. Data are expressed as n (%) unless otherwise specified.

### Primary Outcomes

#### SARS-CoV-2 clearance

The 14-day SARS-CoV-2 clearance rate was 80.0% (12/15) for patients in grp 2 *versus* 16.7% (2/12) for patients in grp 1 (P=0.002). Grp 2 patients had a more rapid clearance of SARS-CoV-2 than in grp 1 (Figure 2). The median time to SARS-CoV-2 clearance in all grp 2 patients that cleared (n=12) was 6.0 days (range 1-12, IQR 1-12), vs only 2 patients clearing virus in grp 1 (statistical analysis not performed secondary to the small number of clearances in grp 1).

**Figure 2.**
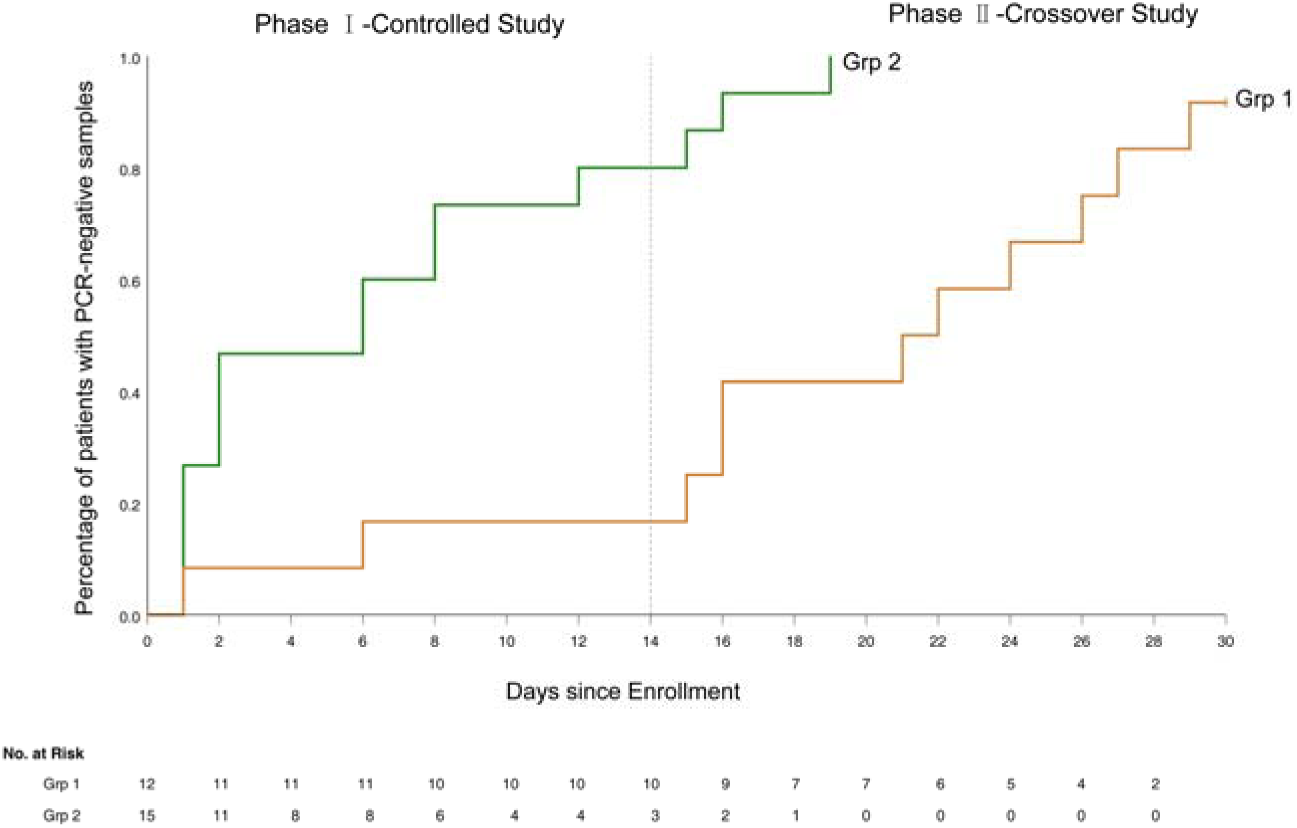
The Kaplan-Meier Curve of Time to SARS-CoV-2 Clearance.

Three patients who remained RT-PCR positive for SARS-CoV-2 despite 2 weeks of leflunomide therapy continued to receive leflunomide and they achieved SARS-CoV-2 clearance on day 15, 16 and 19, respectively. Nine patients receiving standard care remained RT-PCR positive for SARS-CoV-2 on day 14 post-enrollment and were crossed over to receive leflunomide. Eight patients achieved SARS-CoV-2 clearance in a median duration of 9 days (range 0-14, IQR 1-13) from crossover. (Figure 4). By day 30, 15/15 (100%) of patients in grp 2 and 11/12 (91.7%) patients in grp 1 cleared virus. One patient (Pt. 24) in grp 1 initiated leflunomide treatment on day 14 when SARS-CoV-2 tests continued to be positive. After 6 days of leflunomide treatment, SARS-CoV-2 assay converted to negative twice on days 20 and 21. This patient decided to stop leflunomide on day 21. The test of SARS-CoV-2 was positive on day 23 and persisted in positive to day 30 without being discharged.

#### Discharge

The 14-day discharge rate was 73.3% (11/15) for grp 2 *versus* 8.3% (1/12) for grp 1 (*P*=0.001) while the 30-day discharge rate was 100% (15/15) for grp 2 *versus* 66.7% (8/12) patients for grp 1. The median hospital stay was 11 days (IQR 7-19) for grp 2 *versus* 24.0 days for grp 1 (*P*< 0.001) (Figure 3).

**Figure 3.**
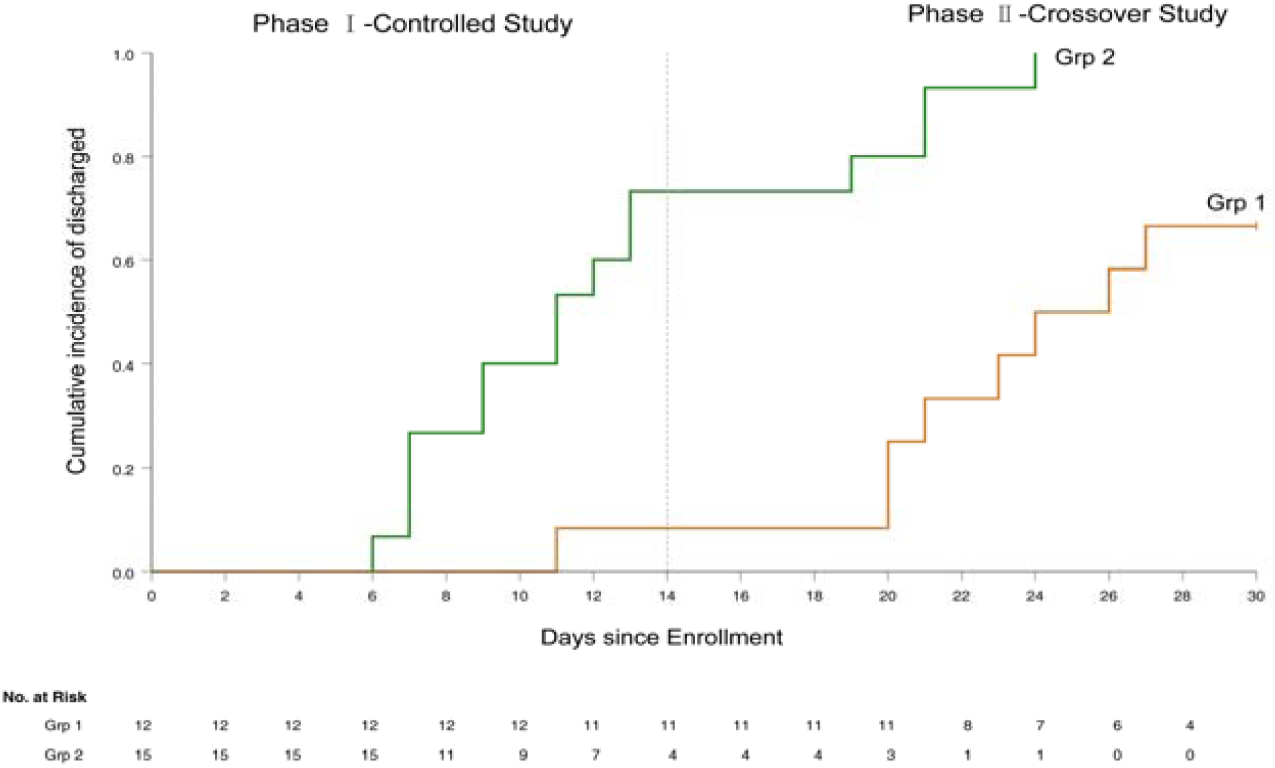
The Kaplan-Meier Curve of Time to Discharge from Hospital.

#### Sensitivity analysis

In order to further confirm the efficacy of leflunomide in enhancing the clearance of SARS-CoV-2, a sensitivity analysis was performed with the elimination of the patients whose viruses were cleared on day one after the enrollments. Based on this principle, four patients in the leflunomide group (Grp 2) and one patient in standard care (grp 1) were eliminated. Therefore, 11 patients left in each group for analysis. The results of the sensitivity analysis showed in Figures S1, S2, and S3. The sensitivity analysis results are consistent with the overall primary endpoint results.

### Secondary Outcomes

#### Flares (SARS-CoV-2 recurrence)

Flare occurred in one patient in group 2 (Figure 4). The patient (Pt. 12) became viral negative on day 1. On day 11, the patient tested positive SARS-CoV-2. The patient tested negative on day 12 and onward, and he was discharged on day 19.

**Figure 4.**
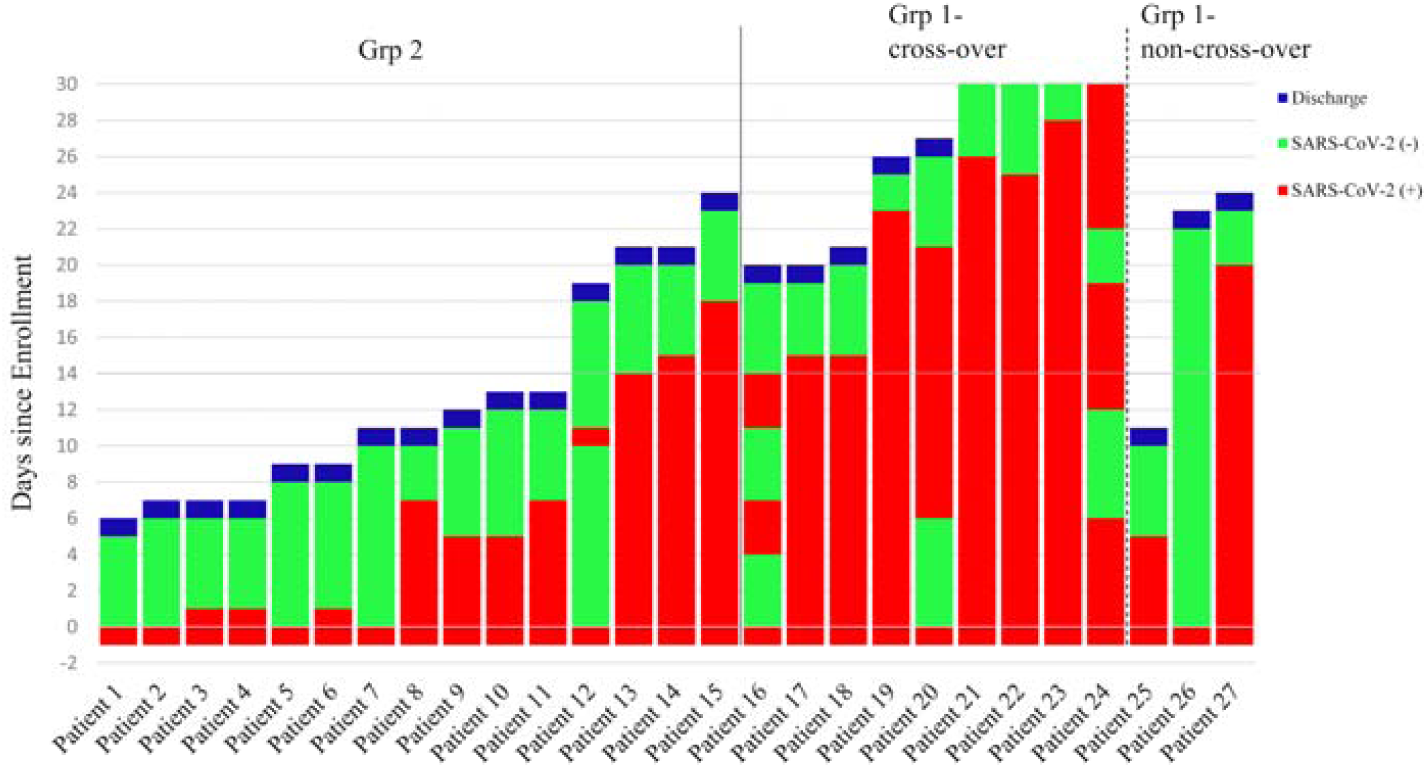
Time Course of Statuses of SARS-CoV-2 Test and End Points. Patients 1 to 15 were in the leflunomide group and patients 16 to 27 were in the standard care group.

### Safety

AEs were observed in 73.3% (11/15) patients in grp 2 and 83.3% (10/12) patients in group 1. Treatment Emergent Adverse Event (TEAEs) were observed in 40% (6/15) patients in grp 2 and 25% (3/12) patients in group 1 (Table S2). The most frequent TEAEs were hyperlipidemia (20%), leukopenia (20%), and neutropenia (13.3%) in the grp 2 and hyperlipidemia (16.7%) and hypoalbuminemia (8.3%) in grp 1. No SAEs or deaths were reported.

## Discussion

This open-label study demonstrated that leflunomide shortened the time to SARS-CoV-2 clearance, improved the SARS-CoV-2 clearance rate and the 14-day hospital discharge rate of COVID-19 patients refractory to standard care, suggesting that DHODH inhibition can provide clinical anti-viral activity and that the FDA approved drugs could be used in COVID-19 patients. Although not the typical viral course, we observed that 2-5% COVID-19 patients fail to clear SARS-CoV-2 as long as 28 days, indicating that a significant proportion of COVID-19 patients may harbor SARS-CoV-2 for an extended period of time^29^. This phenomenon of chronic viral shedding may be a manifestation of relative immune-incompetence related to the SARS-Cov-2 infection, severe initial infection depleting the host’s immune capacity, or underlying immune-dysregulation. The clinical manifestations are usually stabilized with standard care but show persistent SARS-CoV-2 positivity. The clinical and infectious consequences of the continued viral positivity is currently unknown, but we assume that at least a portion of these patients are still contagious and could redevelop clinical symptoms. As they may pose an infectious risk to the community, these patients could not be discharged from the hospital. Recently Remdesivir was granted emergency use authorization from the FDA^30^, however, full approval from the FDA is still several months away. Accelerating the development of vaccines and new drugs is warranted; however, new drug development is time-consuming and cannot meet the immediate and urgent needs. Therefore, repurposing approved medicine is a rational option^7^. Several investigators have looked to currently approved drugs to impact COVID-19 and many have not lived up to expectations^2-6^.

To our knowledge, our study is the first clinical study evaluating a DHODH inhibitor, leflunomide, for the treatment of COVID-19. Leflunomide is the prodrug to the active drug teriflunomide. It has been approved for the treatment of rheumatoid arthritis^9^ by the USA Food and Drug Administration (FDA), lupus nephritis^31^ in China, and psoriatic arthritis^11^ in the EU. Leflunomide is well tolerated^9-10^. Anti-viral activities have been described for leflunomide in CMV^16^ and BK polyomavirus^13-14^ Both the anti-viral and immunosuppressive activities of leflunomide have been attributed to its inhibition of *de novo* pyrimidine synthesis at concentrations active for rheumatoid arthritis^8,12,^, and inhibition of tyrosine and serine kinase activities at higher teriflunomide concentrations^23^. Additionally, previous studies^14,32^ reported that leflunomide inhibits the PI3K-AKT-mTOR pathway, and the release of viral genetic material from capsids depends on this signaling pathway. More recently, *in vitro* studies^24^ indicated that teriflunomide has anti-SARS-CoV-2 activities at a range of 6-26 μM that falls within the recognized therapeutic levels for rheumatoid arthritis. Their data suggested that SARS-CoV-2 was sensitive to teriflunomide *via* inhibition of DHODH^24^, a rate-limiting enzyme in pyrimidine *de novo* synthesis. DHODH catalyzes the dehydrogenation of dihydroorotate to orotic acid to generate uridine and cytosine nucleotides. The authors further speculated that under normal conditions, nucleotides are supplied *via* both *de novo* biosynthesis and the salvage pathway that recycles pre-existing nucleotides from food or other nutrients. The salvage pathway is sufficient for supplying pyrimidines in non-proliferating quiescent cells, but in virus-infected cells, the *de novo* nucleotides biosynthesis is critical to supply the larger intracellular nucleotide pool required for viral replication. Finally, Xiong et al. reported that influenza A virus replication was reduced in cells deficient in *DHODH-/-cells*, even though cell growth is not affected^24^.

In the current study, we investigated leflunomide as additional pharmacologic therapy for hospitalized patients who were RT-PCR-positive for SARS-CoV-2 for more than 28 days despite receiving standard of care. The study showed that leflunomide noticeably increased the 14 days SARS-CoV-2 clearance rate of refractory COVID-19 patients (80.0% *versus* 16.7% for standard care). In addition, leflunomide shortened the median time to SARS-CoV-2 clearance, which was 6.0 days for patients receiving leflunomide, whereas in the standard care group, only two of 12 patients converted to SARS-CoV-2 RT-PCR negative on day 1 and 6. Notably, 9 COVID-19 patients who remained SARS-CoV-2 positive crossed over to leflunomide treatment after two weeks of standard care, 8 patients achieved SARS-CoV-2 clearance approximately 9 days from crossover. In addition, leflunomide significantly increased the 14-day discharge rate (73.3% *versus* 8.3% for standard care) and greatly shortened length of hospital stay. In this study, we applied leflunomide at 20-30 mg/day, matching the dose range for rheumatoid arthritis^9,10^ or systemic lupus erythematosus^30^. We found that leflunomide showed good tolerability and efficacy in treating COVID-19, with an acceptable safety profile of leflunomide, with no SAEs or deaths reported.

### Limitations of this study

Despite the demonstration of promising efficacy of leflunomiude, this study has several limitations. This pilot study was not randomized, the sample size of refractory COVID-19 patients was small, and all patients came from a single center. The RT-PCR assay utilized was semi-quantitative, and no plasma was saved to assay for serum teriflunomide levels.

## Conclusion

This non-randomized, open-label controlled trial has demonstrated that leflunomide has a favorable safety profile and is effective in enhancing SARS-CoV-2 clearance in refractory COVID-19 patients. The tolerability of the 14-28-day course of treatment with leflunomide is acceptable. Based on the efficacy and safety profiles of leflunomide on COVID-19 from this pilot study, a randomized controlled clinical study is warranted.

## Data Availability

After publication, study data will be available on reasonable request to the corresponding author.

## Contributors

QW, HG, Yu-L, and XJ contributed equally to this paper and share joint first authorship. ZH was chief investigator of this study. QW, HG, Yu-L, and XJ were responsible for the design, analysing, and writing of the manuscript. XH, NZ, JF, DS, ZB, and YZ were responsible for recruitment and clinical care of the patients. YH, YS, XY, Yuan-L, BJ, and YG were responsible for the data analyses. Yan-L, FQ, and YW were responsible for the sample collection and laboratory analysis. All authors reviewed and approved the manuscript.

## Funding

This work was supported by the COVID-19 Emergency Tackling Research Project of Shandong University (Grant No. 2020XGA 01).

## Competing interests

All authors have completed the ICMJE uniform disclosure form at www.icmje.org/coi_disclosure.pdf and declare: no support from any organisation for the submitted work other than the listed above; no financial relationships with any organisations that might have an interest in the submitted work in the previous three years; no other relationships or activities that could appear to have influenced the submitted work.

## Ethical approval

The study was approved by Qilu Hospital, Cheeloo College of Medicine, Shandong University (KYLL2020372). All patients gave written informed consent.

## Data sharing

After publication, study data will be available on reasonable request to the corresponding author.

## Dissemination to participants and related patient and public communities

We plan to submit this manuscript to a preprint.

The corresponding author (ZH) affirms that the manuscript is an honest, accurate, and transparent account of the study being reported; that no important aspects of the study have been omitted; and that any discrepancies from the study as planned have been explained.

## Notes

### Competing Interest Statement

The authors have declared no competing interest.

### Clinical Trial

ChiCTR2000033372

